# Evaluation of Resolution-Aware Training Strategies for Deep Learning Detection of Calcaneus Fractures on X-Ray

**DOI:** 10.1101/2025.09.04.25334786

**Authors:** Nicholas J. Yee, Atta Taseh, Samir Ghandour, Evan Sirls, Mansur Halai, Cari Whyne, Christopher W. DiGiovanni, John Y. Kwon, Soheil Ashkani-Esfahani

## Abstract

**Background:** Calcaneus fractures are challenging to identify on radiographs because diagnostically relevant features are subtle and affected by image resolution. Although convolutional neural networks (CNN) have shown strong fracture detection performance, CNN training strategies robust to different image resolutions remain insufficiently characterized.

**Methods:** This retrospective study included foot radiographs from a hospital between 2015 and 2022, comprising 1,775 x-ray series (551 fractures; 1,224 without), split into training (70%), validation (15%), and testing (15%). ImageNet pre-trained ResNet models were fine-tuned on the dataset. Three training strategies were evaluated: (1) single-size training on 128×128, 256×256, 512×512, 640×640, or 900×900 radiographs (five model sets); (2) curriculum learning, sequentially trained from 128×128 to 900×900 (five model sets); and (3) multi-scale augmentation, trained on images continuously resized between 128×128 and 900×900 (one model set). Training and inference times were compared.

**Results:** Multi-scale augmentation achieved the highest average area under the receiver operating characteristic curve (0.938; 95% CI: 0.936–0.939) across image resolutions without increased training or inference time. Curriculum learning demonstrated the highest sensitivity for in-distribution low-resolution images (85.4%–90.1%) and out-of-distribution high-resolution images (78.2%–89.2%) but required significantly longer training times (11.8 [IQR: 11.1–16.4] hours; *P*<.001).

**Conclusions:** While 512×512 images performed well for fracture detection, curriculum learning and multi-scale augmentation improved robustness across image resolutions without additional annotations.

**Summary statement:** Different deep learning training strategies affect performance in detecting calcaneus fractures on radiographs across in- and out-of-distribution image resolutions, with a multi-scale augmentation strategy conferring the greatest overall performance improvement in a single model.

**Key points:**

1. Training strategies addressing differences in radiograph image resolution (or pixel dimensions) could improve deep learning performance.
2. The highest average performance across different image resolutions in a single model was achieved by multi-scale augmentation, where the sampled training dataset is uniformly resized between square resolutions of 128×128 to 900×900.
3. Compared to model training on a single image resolution, sequentially training on increasingly higher resolution images up to 900×900 (i.e., curriculum learning) resulted in higher fracture detection performance on images resolutions between 128×128 and 2048×2048.

## Introduction

Calcaneus fractures are challenging orthopaedic injuries to diagnose on radiographs due to the complex anatomy of the hindfoot and potential for subtle or minimally displaced fracture patterns. Conventional x-rays remain the primary imaging modality in acute settings, yet their diagnostic performance can be limited by the complex hindfoot osteology. Recent developments in using deep learning (DL) to detect fracture patterns on radiographs have demonstrated performance comparable to diagnostic radiologists, suggesting a potential opportunity for an automated radiology support tool in identifying calcaneus fractures [1–3].

The image dimensions of diagnostic radiographs are typically 2000 to 3000 pixels [4]. Radiograph dimensions and their associated resolution depend on the x-ray device quality, usage setting, and imaging configuration. The source-to-object and object-to-detector distances will affect the radiograph resolution. DL model performance is dependent on the radiograph image resolutions where higher resolution images enable the detection of fine-grained features, which can be essential in identifying subtle cortical disruptions in minimally displaced fractures. However, increasing resolution imposes greater computational costs (i.e., graphics processing unit memory) and model training time [5–7].

Sabottke and Spieler found that the diagnostic performance of convolutional neural networks (CNN), which is a popular DL architecture that hierarchically extracts spatial features from input images by applying a series of sliding kernels, generally improves with increasing resolution with diminishing returns beyond 448×448 pixels for thoracic pathologies [5–7]. Low-resolution images obscure the details, limiting diagnostic interpretation. To our knowledge, there are no studies reporting the optimal image resolution or exploring image resolution-aware training strategies for CNN-based fracture identification on radiographs.

Given the anatomical complexity of the hindfoot and the diagnostic challenge posed by subtle calcaneus fractures, DL algorithms may enable diagnosis with higher accuracy and consistency. Furthermore, investigating methods to improve the performance of DL training algorithms could bring about a significant added value. In this study, we hypothesized that there are optimal training strategies that address the diversity of image resolutions to improve DL models in identifying calcaneus fractures. The purpose of this study is to investigate three training strategies to balance the trade-offs between the model performance across image resolutions and training/inference computational resources.

## Materials and Methods

This was a retrospective study conducted at a single academic medical center. Institutional review board approval was received (IRB no. 2015P000464). Anonymized diagnostic radiology imaging data were collected from the institution’s imaging archive between 2015 and 2022. A total of 1,775 complete foot X-ray series (AP, oblique, and lateral) were included: 551 series with a calcaneus fracture and 1,224 without a fracture. Radiographs were viewed using the bone window and without overlays. Females represented 43% of the patients (60 ± 17 years old) and males were 57% (53 ± 15 years old). The foot radiograph dimensions were 1942 ± 992 pixels in height by 2114 ± 767 pixels in width.

The classification of fracture status was based on the clinical radiology reports by board-certified diagnostic radiologists. When radiographs were equivocal for fractures, the CT report was used to confirm the fracture status. DL models were pre-trained on ImageNet and fine-tuned on the calcaneus training dataset [8]. The dataset was partitioned randomly by image series (70% training, 15% validation, 15% test). Training image augmentation included random horizontal flipping (probability, P = 0.5); random rotation up to ±90° (P = 0.25); random affine transformation including rotation (±10°), scaling (0.9–1.1×), translation (up to 5% in both axes), and shear (up to 5°) (P = 0.3); Gaussian noise addition with mean 0.0 and standard deviation 0.1 (P = 0.3); and color jittering adjusting brightness, contrast, and saturation (±0.2) and hue (±0.1) (P = 0.3).

A model set included ResNet-18, −34, −50, and −101 and was trained using PyTorch (version 2.4.1+cu121; https://pytorch.org/) and TorchVision (version 0.19.1+cu121; https://docs.pytorch.org/vision) on either a NVIDIA RTX 4090 or the high-performance computing cluster (HPC) with NVIDIA Tesla V100 graphics processing units (NVIDIA, California, USA). The Python version used was 3.10.16 distributed by conda-forge (https://anaconda.org/conda-forge/python). Model weights initialization used TorchVision ImageNet v2 weights for ResNet-50 and −101 and used v1 weights for ResNet-18 and −34. Inputs used the AP, lateral, and oblique views as a 3-channel image. The radiographs were normalized following the intensity distribution of the training dataset to stabilize training curves without introducing data leakage. Subsequently, they were zero-padded symmetrically along its short side to square dimensions before bilinear interpolation to the target dimensions. Symmetric padding maintained the native aspect ratio and avoided geometric distortion of fracture morphology. The final classification layer was replaced with a He-initialized multilayer perceptron for binary classification. Models were trained end-to-end with no frozen layers using binary cross entropy loss with weight decay, label smoothing, and early stopping.

Three training strategies were investigated: (1) single size images, where 5 sets of ResNet models were trained on images resized to 128×128, 256×256, 512×512, 640×640, or 900×900; (2) curriculum learning, where the models were progressively trained from lower to higher image resolutions through image downsizing with bilinear interpolation to gradually increase the number of input features (i.e., trained with images with resolutions 128×128 then 256×256 then 512×512 then 640×640 and finally with 900×900; the weights from each completed stage initialized the next stage); and (3) multi-scale augmentation, where models were trained using randomly resized square images between 128×128 and 900×900 to achieve partial scale invariance (**Figure 1**). The detailed training schedule is included (Supplemental Table 4). All models were trained with 15 random hyperparameter sets with the best hyperparameters selected using the highest area under the Receiver Operating Characteristic curve (AUROC) on the validation dataset. The hyperparameter sets sampled from predefined ranges (Supplemental Table 4). Batch size was optimized based on model architecture and image resolutions (Supplemental Table 5).

**Figure 1:**
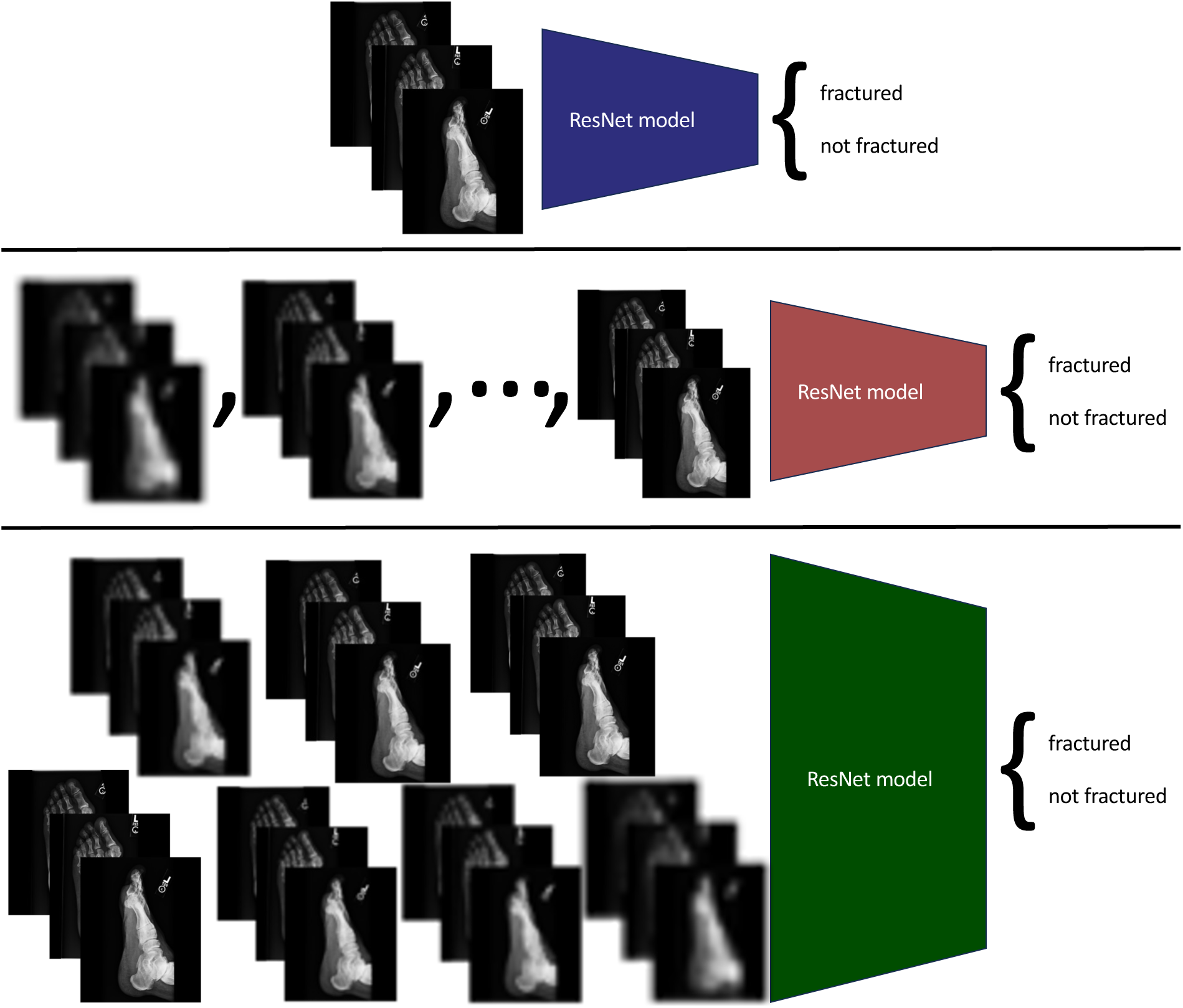
Resolution-aware training strategies for calcaneus fracture detection. Overview of the training strategies for calcaneus fracture detection using ResNet models (top: single size images; middle: curriculum learning sequentially from low to high image resolutions; and bottom: multi-scale augmentation).

Fracture detection performance on the held-out test dataset was reported by averaging the ResNet models’ sensitivity/recall, specificity, and AUROC using 500 iterations of bootstrapping. For each bootstrapping iteration, a half-sized test subset was randomly sampled with replacement from the held-out test dataset. Inference time per image across ResNet architectures and image resolutions was calculated using a batch size of one on the workstation equipped with a NVIDIA RTX 4090. The training times were reported for the different training strategies on the HPC using a single NVIDIA V100 and compared using a Kruskal-Wallis test with post-hoc Dunn’s test.

## Results

We assessed the average performance of the DL models on the test images padded and resized to the square dimensions 128×128, 256×256, 512×512, 640×640, 900×900, 1024×1024, and 2048×2048. When the performance was averaged across all those image resolutions, models trained using multi-scale augmentation achieved the highest average AUROC of 0.938 [95% CI: 0.936 - 0.939] compared to the other training strategies (Figure 2; Table 1). Among the curriculum learning models, the model set trained on images up to 512×512 had the highest average AUROC of 0.914 [0.913 - 0.915]. The models trained with the single size strategy on images of 512×512 or 640×640 had the highest average AUROC of 0.903 [0.901 - 0.905] and 0.903 [0.900 - 0.905], respectively.

**Figure 2:**
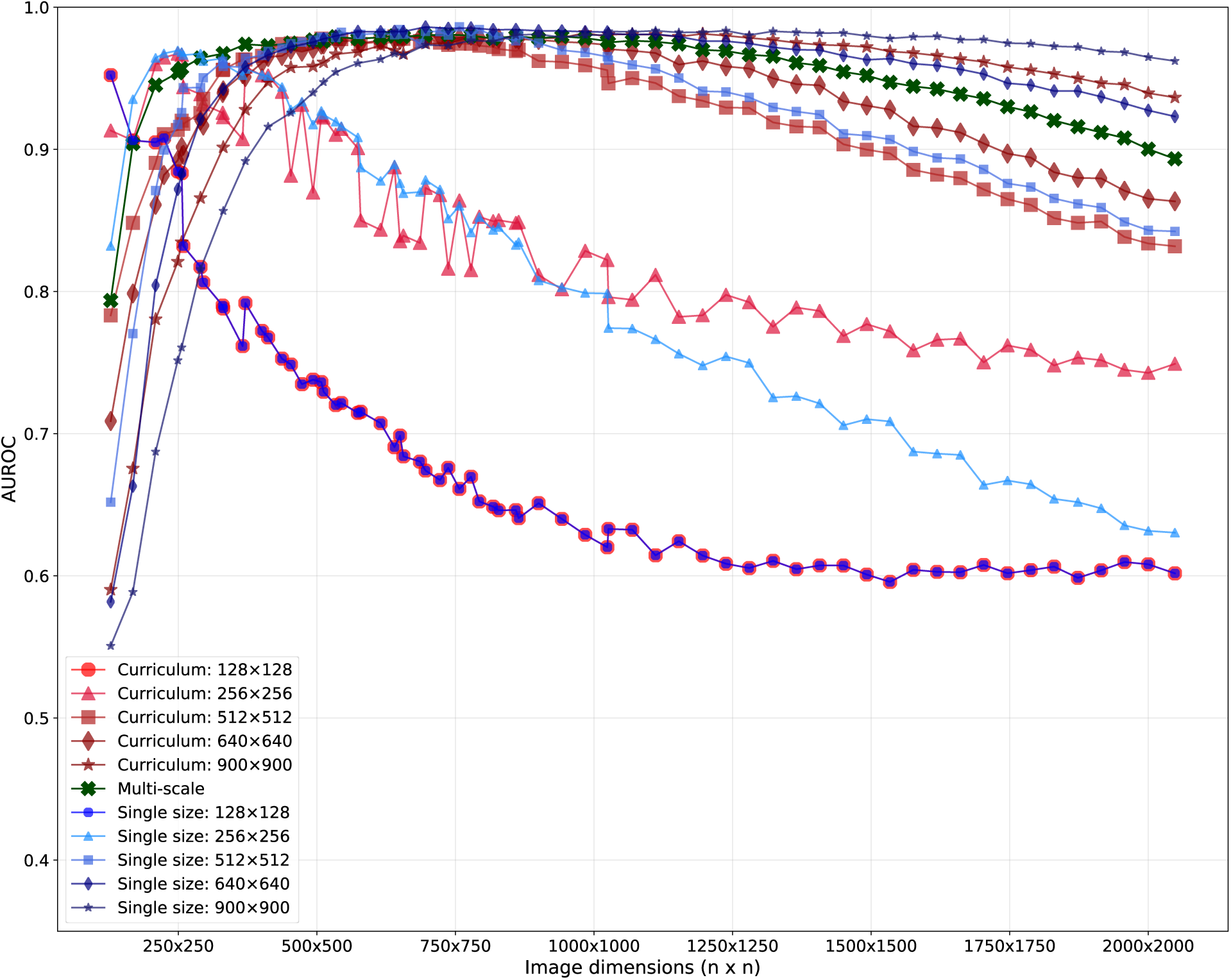
AUROC across image resolutions for different training strategies. Average AUROC of the ResNet models trained under curriculum learning, multi-scale augmentation, and single size training strategies across image dimensions (128×128 to 2000×2000). Curriculum training started on 256×256 images and the weights initialized using the single size 128×128 models (represented by the blue dot with a red outline).

**Table 1:** AUROC of the training strategies trained on different image resolutions. Reports the average AUROC of the ResNet-18, −34, −50, and −101 models on a held-out test dataset. Low resolution includes square image resolutions between 128×128 to 900×900. High resolution includes image resolutions 1024×1024 and 2048×2048. Bolded values indicate best performance.

| Test image dimensions: |  | 128x128 | 256x256 | 512x512 | 640x640 | 900x900 | 1024x1024 | 2048x2048 | Low resolution | High resolution |
| --- | --- | --- | --- | --- | --- | --- | --- | --- | --- | --- |
| Training strategy | Training image dimension | AUROC [95% CI] | AUROC [95% CI] | AUROC [95% CI] | AUROC [95% CI] | AUROC [95% CI] | AUROC [95% CI] | AUROC [95% CI] | Average AUROC [95% CI] | Average AUROC [95% CI] |
| Curriculum learning | 128x128 | N/A | N/A | N/A | N/A | N/A | N/A | N/A | N/A | N/A |
| Curriculum learning | Up to 256x256 | 0.907<br>[0.906–0.908] | 0.967<br>[0.966–0.967] | 0.926<br>[0.925–0.927] | 0.887<br>[0.885–0.888] | 0.808<br>[0.804–0.812] | 0.826<br>[0.824–0.829] | 0.745<br>[0.742–0.748] | 0.899<br>[0.898–0.900] | 0.786<br>[0.783–0.788] |
| Curriculum learning | Up to 512x512 | 0.775<br>[0.773–0.778] | 0.919<br>[0.918–0.921] | 0.976<br>[0.976–0.976] | 0.981<br>[0.981–0.981] | 0.963<br>[0.963–0.964] | 0.955<br>[0.955–0.956] | 0.828<br>[0.827–0.829] | 0.923<br>[0.921–0.925] | 0.892<br>[0.890–0.894] |
| Curriculum learning | Up to 640x640 | 0.713<br>[0.710–0.716] | 0.914<br>[0.912–0.915] | 0.975<br>[0.975–0.975] | 0.976<br>[0.975–0.976] | 0.976<br>[0.976–0.977] | 0.972<br>[0.971–0.972] | 0.861<br>[0.859–0.862] | 0.911<br>[0.909–0.913] | 0.916<br>[0.914–0.918] |
| Curriculum learning | Up to 900x900 | 0.589<br>[0.585–0.592] | 0.834<br>[0.831–0.836] | 0.960<br>[0.960–0.961] | 0.972<br>[0.971–0.972] | 0.980<br>[0.980–0.981] | 0.979<br>[0.979–0.979] | 0.931<br>[0.930–0.932] | 0.867<br>[0.864–0.870] | 0.955<br>[0.954–0.956] |
| Multi-scale | N/A | 0.798<br>[0.795–0.800] | 0.960<br>[0.959–0.960] | 0.978<br>[0.977–0.978] | 0.980<br>[0.980–0.980] | 0.980<br>[0.979–0.980] | 0.979<br>[0.978–0.979] | 0.893<br>[0.891–0.894] | **0.939**<br>**[0.937–0.940]** | 0.936<br>[0.934–0.937] |
| Single size | 128x128 | **0.954**<br>**[0.953–0.955]** | 0.887<br>[0.886–0.889] | 0.733<br>[0.729–0.737] | 0.696<br>[0.694–0.699] | 0.671<br>[0.670–0.673] | 0.619<br>[0.617–0.621] | 0.604<br>[0.599–0.609] | 0.788<br>[0.786–0.791] | 0.612<br>[0.609–0.614] |
| Single size | 256x256 | 0.829<br>[0.825–0.833] | **0.970**<br>**[0.970–0.971]** | 0.925<br>[0.924–0.926] | 0.896<br>[0.895–0.898] | 0.801<br>[0.797–0.806] | 0.801<br>[0.798–0.805] | 0.631<br>[0.626–0.635] | 0.884<br>[0.883–0.886] | 0.716<br>[0.712–0.720] |
| Single size | 512x512 | 0.664<br>[0.661–0.667] | 0.920<br>[0.919–0.922] | 0.979<br>[0.979–0.979] | 0.978<br>[0.978–0.979] | 0.974<br>[0.973–0.974] | 0.967<br>[0.966–0.967] | 0.837<br>[0.834–0.839] | 0.903<br>[0.901–0.906] | 0.902<br>[0.899–0.904] |
| Single size | 640x640 | 0.580<br>[0.576–0.584] | 0.886<br>[0.885–0.888] | **0.981**<br>**[0.981–0.982]** | **0.985**<br>**[0.985–0.986]** | **0.983**<br>**[0.983–0.984]** | **0.983**<br>**[0.982–0.983]** | 0.922<br>[0.921–0.923] | 0.883<br>[0.880–0.886] | 0.952<br>[0.951–0.953] |
| Single size | 900x900 | 0.555<br>[0.553–0.557] | 0.755<br>[0.752–0.758] | 0.947<br>[0.946–0.947] | 0.967<br>[0.966–0.967] | 0.982<br>[0.981–0.982] | 0.980<br>[0.979–0.980] | **0.959**<br>**[0.958–0.959]** | 0.841<br>[0.838–0.844] | **0.969**<br>**[0.969–0.970]** |

Across the in-distribution test resolutions (128×128 - 900×900), the highest AUROC at each resolution was achieved by the single size models trained at or near that resolution (Table 1). The AUROC of the curriculum learning and multi-scale augmentation models was typically within 0.01 of the best. For the out-of-distribution high-resolution images, all three strategies remained strong at 1024×1024 (AUROC ≥ 0.979). At 2048×2048, all strategies decreased in AUROC, with single size training on 900×900 images retaining the highest AUROC (0.959 [0.958 - 0.959]) compared to multi-scale augmentation models (0.893 [0.891 - 0.894]) and curriculum learning models (0.931 [0.930 - 0.932]).

On test images of resolution from 256×256 to 900×900, the sensitivity of the best model sets trained under curriculum learning, multi-scale augmentation, and single size strategies ranged between 85.4% to 90.1%, 82.8% to 87.7%, and 84.2% to 87.5%, respectively (Table 2). Their specificity was between 98.4% to 99.4%, 96.4% to 98.9%, and 98.7% to 99.3%, respectively. The sensitivity when evaluated on the out-of-distribution high-resolution 1024×1024 and 2048×2048 test images was 89.2 [89.1–89.4] and 78.2 [78.0–78.4] for curriculum learning models, 79.7 [79.4–80.1] and 49.4 [48.7–50.1] for multi-scale augmentation models, and 84.5 [84.3–84.6] and 72.8 [72.4–73.1] for single size trained models, respectively (Table 3). The average of the ResNet models trained under the curriculum learning strategy consistently achieved the highest sensitivity across all image resolutions. The ResNet models trained with multi-scale augmentation had a drastic drop in sensitivity on the 2048×2048 images compared to the lower resolution images.

**Table 2:**
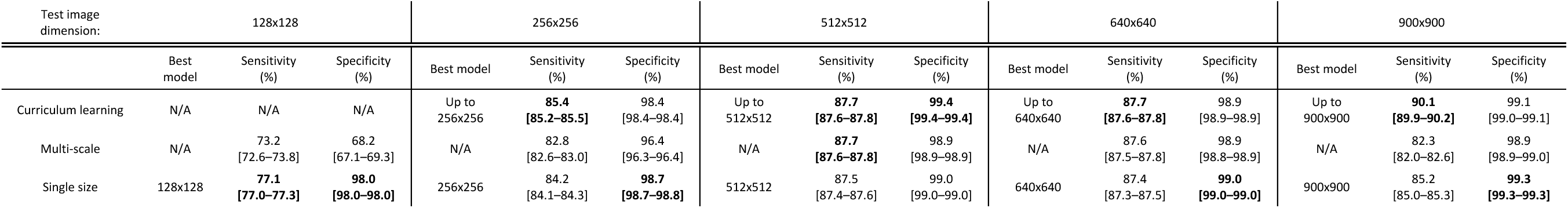
Sensitivity and specificity with their 95% confidence interval on a held-out test dataset resized to different image resolutions. The bolded numbers indicate the best performance for that image dimension. The highest sensitivity was consistently achieved by ResNet models trained using curriculum learning. The best performance among the curriculum learning and single size trained models was reported. The same multi-scale trained models were used for all test image dimensions.

**Table 3:** Sensitivity and specificity with their 95% confidence interval on a held-out test dataset resized to out-of-distribution image dimensions. The bolded numbers indicate the best performance for that image dimension. The highest sensitivity was consistently achieved by ResNet models trained using curriculum learning. The best performance among the curriculum learning and single size trained models was reported. The same multi-scale trained models were used for all test image dimensions.

| Test image dimension: | 1024x1024 |  |  | 2048x2048 |  |  |
| --- | --- | --- | --- | --- | --- | --- |
|  | Best model | Sensitivity (%) | Specificity (%) | Best model | Sensitivity (%) | Specificity (%) |
| Curriculum learning | Up to 900x900 | **89.2 [89.1–89.4]** | 99.1 [99.1–99.2] | Up to 900x900 | **78.2 [78.0–78.4]** | 95.5 [95.5–95.6] |
| Multi-scale | N/A | 79.7 [79.4–80.1] | 98.7 [98.7–98.8] | N/A | 49.4 [48.7–50.1] | 98.3 [98.3–98.4] |
| Single size | 900x900 | 84.5 [84.3–84.6] | **99.3 [99.3–99.4]** | 900x900 | 72.8 [72.4–73.1] | **99.2 [99.1–99.2]** |

Generally, curriculum learning trained models performed the best on test images that matched the image resolution of the final training image resolution with the highest sensitivity continuously increasing up to 900×900 images. The best single size trained models performed well on image resolutions centered around the training image resolution and, out of all the single sized trained models, the best sensitivity was on 512×512 images. Models trained with multi-scale augmentation had the best performance on test images centered around 512×512 (Figure 2 and Figure 3).

**Figure 3:**
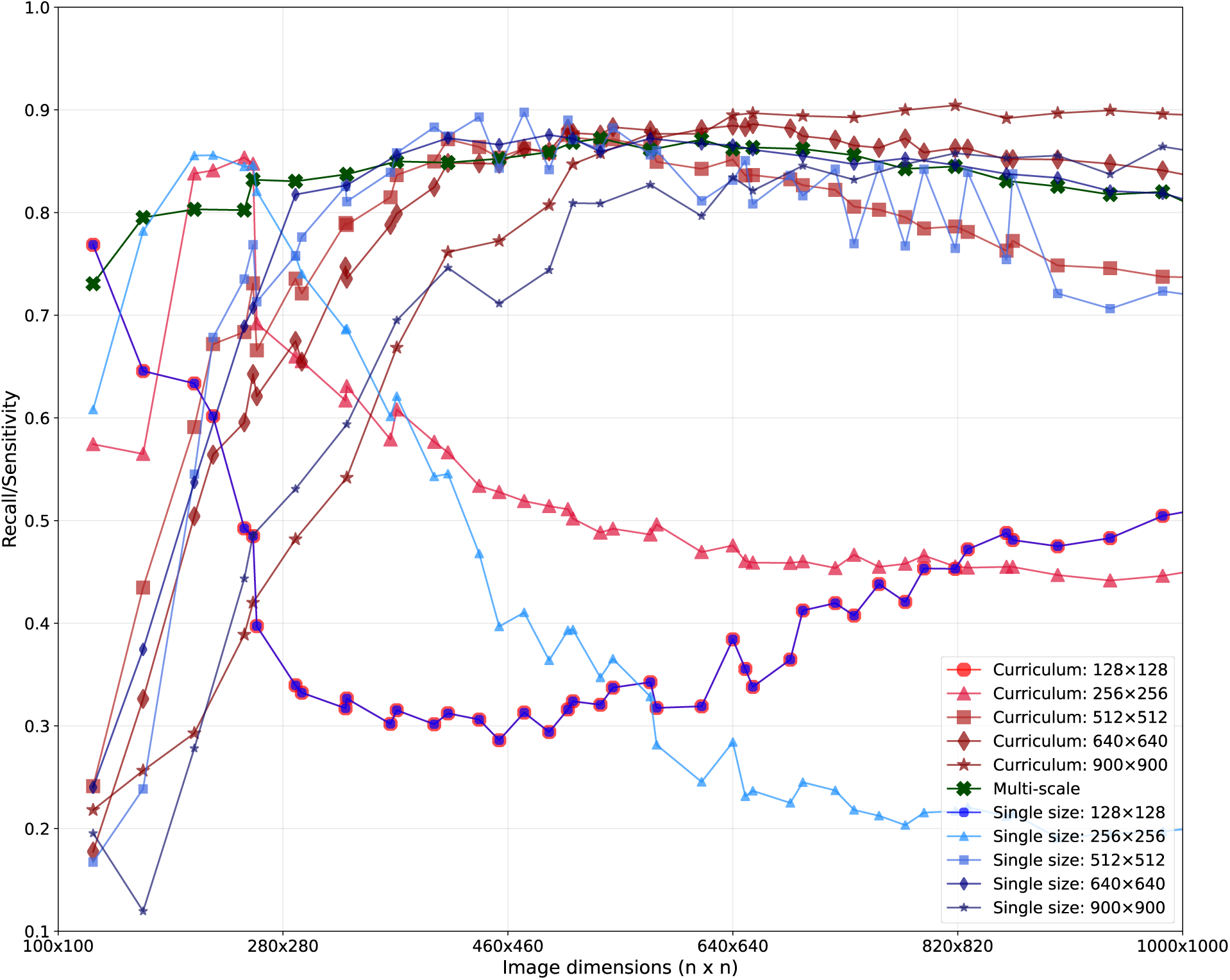
Recall/sensitivity across image resolutions for different training strategies. Average recall/sensitivity of the ResNet models trained under curriculum learning, multi-scale augmentation, and single size training strategies across image dimensions (128×128 to 900×900). Curriculum training started on 256×256 images and the weights initialized using the single size 128×128 models (represented by the blue dot with a red outline).

Curriculum learning models took significantly longer to train (11.8 [interquartile range: 11.1–16.4] hours) than multi-scale (6.7 [4.9–8.9] hours) and single size (8.0 [5.6–10.4] hours) training strategies (*P*<.001; Supplemental Table 4). The inference speed using the smallest CNN model, ResNet-18, was 4.59 times faster than the largest model, ResNet-101 (Figure 4). Inference speed was 1.66 times faster using 128×128 images than 1024×1024 images.

**Figure 4:**
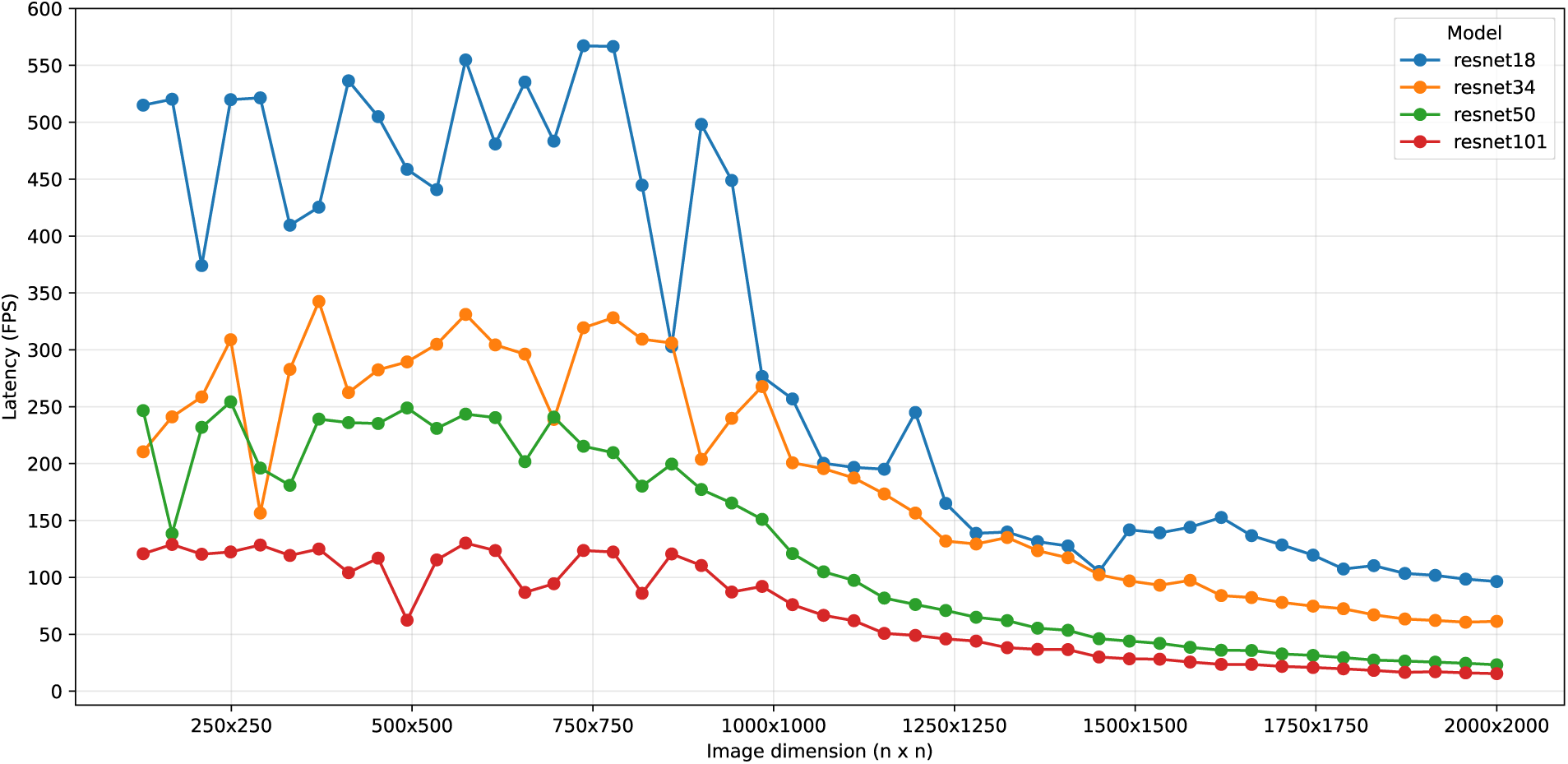
Inference speed across image resolutions on a RTX 4090. Inference time (frames per second; FPS) on an NVIDIA RTX4090 with a batch size 1 for each ResNet model at different image dimensions.

## Discussion

### Comparison of model performance between training strategies

Prior work on assessing the impact of radiograph image resolution on DL classification performance focused on chest radiographs and mammography [5–7,9–11]. Sabottke and Spieler and Wollek et al. both evaluated training at different fixed input resolutions on chest radiographs, including 32×32 dimensions, which calcaneus fracture features could not be reliably identified on a radiograph in clinical practice [7,9]. Wollek et al. reported the highest classification performance at 1024×1024 on chest radiographs, motivating our extension of the inference evaluation to 2048×2048. Niu et al. take a structurally different approach to multi-scale interpretation on mammograms [12]. Rather than varying the global input resolution, they fuse features from a global patch with those of a lesion-centered local patch, interpreting the image at two scales through an architectural prior. Their approach would require additional bounding box or segmentation annotation, which can be costly to obtain at scale for medical imaging. Our study uniquely compares different training strategies (single size, curriculum learning, and multi-scale augmentation) head-to-head on a shared backbone using only image-level labels.

While DL has been studied in fracture identification on radiographs, to our knowledge, no previous studies have investigated the impact of image resolution on the performance of DL models for fracture identification on radiographs [13–16]. Calcaneus fractures can be difficult to identify on radiographs with a reported sensitivity between 80-100% and specificity between 30-100% [17,18]. On foot and ankle radiographs, diagnosis of fractures has a sensitivity between 66-100% and specificity between 40-100% [17–21]. Our study aims to report the performance of calcaneus fracture identification on radiographs using DL models with different training strategies to overcome challenges with variable radiograph image resolutions. The ResNet models trained with multi-scale augmentation between 128×128 and 900×900 had a high sensitivity and specificity across the range of image resolutions between 256×256 and 1024×1024 with notable weakness with 128×128 and 2048×2048 images. On any image resolution, the ResNet models trained with curriculum learning had the highest sensitivity and specificity, but this strategy required longer training times than the other training strategies. Furthermore, it would need different weights trained up to the desired image resolution for each image resolution as the performance quickly degrades at large mismatches between the training and test image resolutions. Training using a single image size would circumvent the longer training times and would have sensitivity slightly lower than the curriculum learning trained models, even on high-resolution radiographs up to 2048×2048. It shares the same limitation with performance degradation on images resolutions out-of-distribution.

We demonstrated that the model training strategy addressing image resolution resulted in different performance on identifying calcaneus fractures. Bengio et al. first described curriculum learning as a strategy related to boosting algorithms that starts with easier examples and progressively shifts to challenging examples rather than learning on a uniform distribution as is done with multi-scale augmentation [22]. Similar applications of curriculum learning in radiographic fracture detection demonstrated improved performance [23,24]. Their curriculums were based on clinical difficulty graded by clinicians rather than image resolution. Curriculum learning mimics graduated human learning, training the model to represent increasingly complex radiographic features in stages. One interpretation is that each higher-resolution stage mainly refines features already learned at lower resolutions. Consistent with this, the later high-resolution stages trained faster than single-size models built from scratch at the same resolution.

Using the single size training strategy, the sensitivity increased as the image resolution increased up to 512×512 but trended downwards as training image resolution approached 900×900. Other DL approaches to diagnostic imaging interpretation found similar plateauing effects [5–7]. The CNN architecture is designed with hierarchical feature representations with deeper layers capturing a broader receptive field (i.e. a larger subset of the original image is represented in an element of the layer’s feature map) and accumulation of semantic information [25–27]. A smaller anatomical region of the original image is represented at the same depth in a higher resolution image such as 2048×2048 compared to a 512×512 image, resulting in potentially important anatomical landmarks in identifying fractures being omitted. At low resolutions, the radiographic features of a fracture may be indistinguishable, making for poor model image interpretation. The ResNet models trained under multi-scale augmentation appear to follow a similar trend to single size trained models of maximizing sensitivity around 512×512 images. This may reflect the models learning the scale at which fracture features are best resolved, while avoiding noise from unnecessary detail at higher resolutions. Alternatively, the models may be implicitly representing the central range of resolutions, leading to the observed suboptimal performance relative to the curriculum learning and single size trained models at the smallest and largest resolutions.

In contrast to single size and multi-scale training strategies, the models trained with curriculum learning achieved the best sensitivity at each image resolution out of the 3 training strategies and the performance continued to improve up to 900×900 resolution. On out-of-distribution image resolutions, the performance expectedly decreased but still outperformed the models trained with multi-scale augmentation and single size training strategies. While it takes longer to train a model with curriculum learning, the inference time was equivalent as the underlying architecture was the same. The downside would be the additional memory requirements for multiple weights and the inference pipeline would need to route the data towards the appropriate model instance based on the image resolution. A data pipeline requiring maximal performance with ample training compute resources should consider employing a curriculum learning training strategy. The models trained with multi-scale augmentation offer a single model solution for systems with limited compute resources.

### Clinical translation and implications

A resolution-robust decision support tool for interpreting calcaneus radiographs has the potential to meaningfully improve clinical and diagnostic workflows. It can be used as a quality assurance tool by providing a second read. In practice, the model could output the probability of a fracture and a heatmap localizing the fracture alongside the radiograph. In emergency settings, this can reduce missed calcaneus fractures and improve diagnostic confidence of less experienced clinicians. In radiology departments, it could support workflow prioritization by triaging calcaneus imaging suspicious of fractures (that may require arranging a CT scan). Each training strategy yields models suitable for different clinical cases. For example, a mobile fluoroscopy unit in the trauma bay or on sports fields may produce non-standard resolutions. The device’s computer may not have enough resources to evaluate multiple models. A single model trained using multi-scale augmentation may be most efficient. In contrast, the radiology department would have access to hospital servers with sufficient compute to run several models to achieve the highest performance. From a clinical research or implementation perspective, machine learning engineers should consider strategies to address image resolution given their available resources as the diagnostic performance will likely drop off outside the training distribution.

Our aim was to characterize the effect of resolution-aware training strategies on a clear, learnable task where the supervisory signal was well defined. Calcaneus fractures are clinically heterogeneous and span at least four radiographically distinct phenotypes: intraarticular (Sanders I–IV), extraarticular tuberosity avulsion, anterior process, and stress fractures. They can differ markedly in radiographic conspicuity, with stress and extraarticular fractures the most subtle. Fracture subtype classification is a harder signal to learn than fracture detection because within subtype variations can be substantial and certain presentations may border between multiple subtypes. From a labelling perspective, this would likely lead to more disagreements between radiologists or surgeons reading the images for subtype classification compared to detecting fractures alone. Furthermore, certain subtypes will dominate the cohort by frequency and can bias the learned representation toward the more common patterns. Future studies investigating calcaneus fracture subtype classification should collect CT scans for ground truth labels as the Sanders classification for intraarticular fractures is a CT-based system. DL models that can accurately classify intraarticular subtypes on radiographs may be able to influence clinical decision making on the need for advanced imaging.

### Limitations and future directions

Our work focused on assessing training strategies to address the range of image resolution of foot radiograph concerning for calcaneus fractures to improve DL image classification performance. Fundamentally, our work described alternative pathways to explore the model hypothesis space for the optimal weights that minimize our binary classification loss function. Among the countless methods to optimize this non-convex optimization problem, further work in improving robustness towards image resolutions and general fracture identification performance should investigate model pre-training strategies that adapt to challenges faced in the musculoskeletal domain. Our models had their weights initialized through pre-training on the ImageNet dataset containing 14,197,122 images of a variety of image resolutions [8]. This process, known as transfer learning, leverages models trained on larger datasets to improve the performance on tasks with limited data by adapting the learned image representations (i.e. image feature extractors) of general images sourced from the internet towards our target domain of fracture identification on radiographs [28–33]. Further modeling improvements may benefit from domain-adaptive pre-training where model weights are fine-tuned on a larger dataset of musculoskeletal images (which may or may not contain the foot) prior to the final training on foot radiographs [34–36]. FracAtlas and MURA are large public musculoskeletal fracture datasets that present an opportunity for domain-adaptive pre-training to improve weight initialization on downstream tasks [37,38].

As the musculoskeletal radiograph datasets develop in quality and quantity, algorithmic advancements will independently improve model performance. Our results on the model training strategies of curriculum learning and multi-scale augmentation are limited to the foot radiographs collected from a single hospital system for the assessment of a single binary classification task of fracture identification. Our ground truth labels were obtained from the radiograph reports and, if needed, from the CT reports.

Further validation of these training strategies in calcaneus fractures should include multiple institutions and systematic expert image review. Future work should explore the generalizability of these training strategies to other pathologies, external calcaneus datasets, and broader DL tasks including multiclass classification, object detection, segmentation, and keypoint detection. Hyperparameter tuning was performed within the limits of the available computational resources. As hardware advances, there will be opportunities to investigate the effects of larger models, larger batch sizes, and other hyperparameters on a broader range of image resolutions. Additionally, training and inference metrics were reported on the hardware available and training efficiency was optimized to the best of our knowledge and available evaluation period. The image resolution robustness of these training strategies and its generalizability beyond calcaneus radiographs should be validated at external institutions, vendors, and acquisition protocols.

This study demonstrated that DL could identify calcaneus fractures on radiographs of a variety of image resolutions. Furthermore, it compared different training strategies on the performance on in- and out-of-distribution image resolutions and examined the differences in required computational resources. Model training strategies, such as curriculum learning and multi-scale augmentation, present an opportunity to potentially improve model robustness towards different image resolutions without requiring additional annotated data.

## Data Availability

All data produced in the present study are available upon reasonable request to the authors

## Supplemental

**Table 4:**
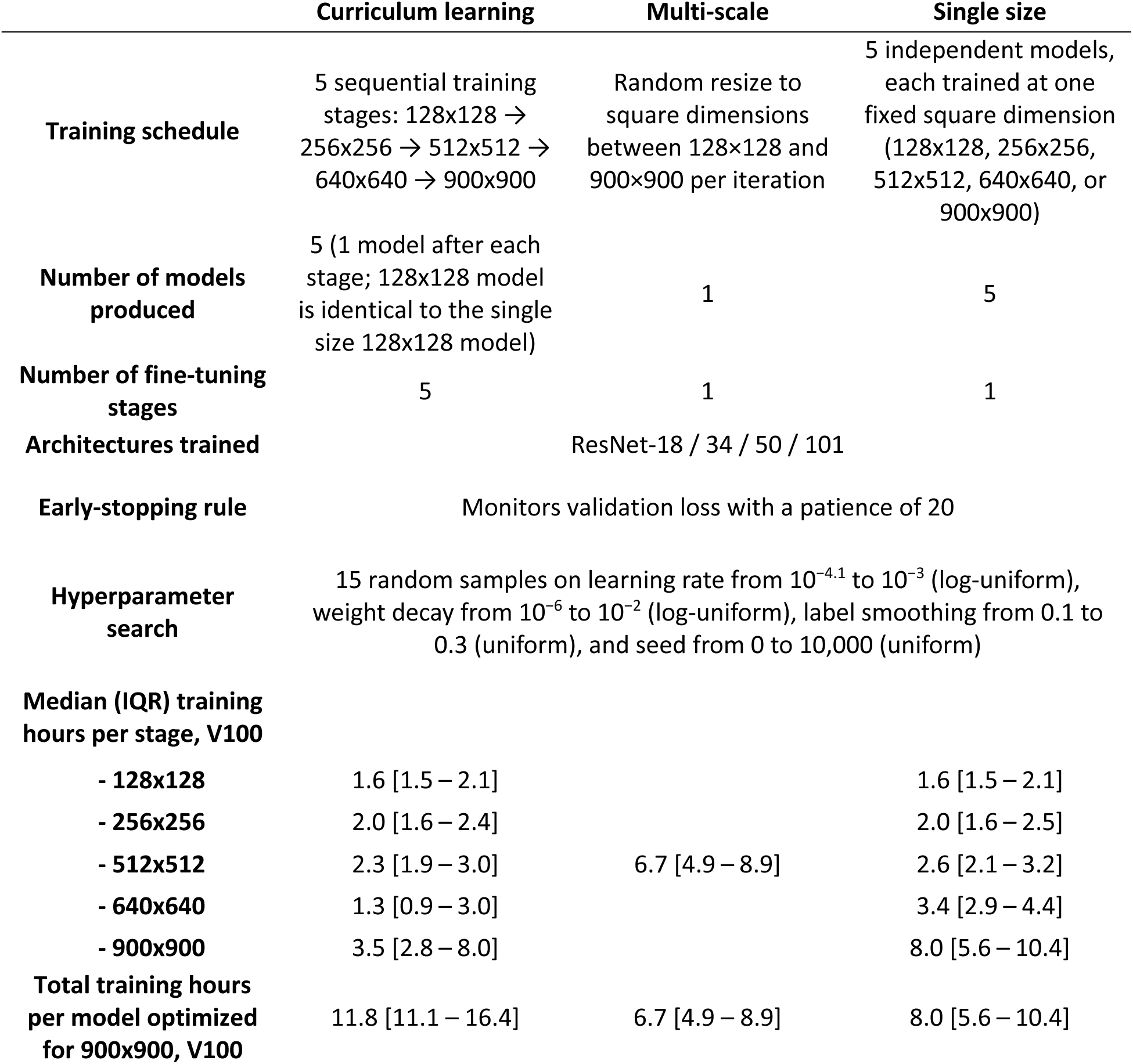
Training schedule and median (interquartile range) training hours per stage for each training strategy on a NVIDIA Tesla V100, pooled across the ResNet-18, −34, −50, and −101 architectures.

|  | Curriculum learning | Multi-scale | Single size |
| --- | --- | --- | --- |
| Training schedule | 5 sequential training stages: 128x128 → 256x256 → 512x512 → 640x640 → 900x900 | Random resize to square dimensions between 128×128 and 900×900 per iteration | 5 independent models, each trained at one fixed square dimension (128x128, 256x256, 512x512, 640x640, or 900x900) |
| Number of models produced | 5 (1 model after each stage; 128x128 model is identical to the single size 128x128 model) | 1 | 5 |
| Number of fine-tuning stages | 5 | 1 | 1 |
| Architectures trained | ResNet-18 / 34 / 50 / 101 |  |  |
| Early-stopping rule | Monitors validation loss with a patience of 20 |  |  |
| Hyperparameter search | 15 random samples on learning rate from 10<sup>-4.1</sup> to 10<sup>-3</sup> (log-uniform), weight decay from 10<sup>-6</sup> to 10<sup>-2</sup> (log-uniform), label smoothing from 0.1 to 0.3 (uniform), and seed from 0 to 10,000 (uniform) |  |  |
| Median (IQR) training hours per stage, V100 |  |  |  |
| - 128x128 | 1.6 [1.5 – 2.1] |  | 1.6 [1.5 – 2.1] |
| - 256x256 | 2.0 [1.6 – 2.4] |  | 2.0 [1.6 – 2.5] |
| - 512x512 | 2.3 [1.9 – 3.0] | 6.7 [4.9 – 8.9] | 2.6 [2.1 – 3.2] |
| - 640x640 | 1.3 [0.9 – 3.0] |  | 3.4 [2.9 – 4.4] |
| - 900x900 | 3.5 [2.8 – 8.0] |  | 8.0 [5.6 – 10.4] |
| Total training hours per model optimized for 900x900, V100 | 11.8 [11.1 – 16.4] | 6.7 [4.9 – 8.9] | 8.0 [5.6 – 10.4] |

**Table 5:** Training batch sizes for each model architecture and input image resolutions.

| Model | Batch size |  |  |  |  |
| --- | --- | --- | --- | --- | --- |
|  | 128x128 | 256x256 | 512x512 | 640x640 | 900x900 |
| ResNet-18 | 256 | 128 | 64 | 64 | 16 |
| ResNet-34 | 256 | 128 | 64 | 64 | 16 |
| ResNet-50 | 256 | 128 | 64 | 32 | 16 |
| ResNet-101 | 256 | 128 | 32 | 32 | 16 |

